# Alzheimer Disease Plasma Biomarkers in the Mid-Western Amish

**DOI:** 10.1101/2024.12.23.24319579

**Authors:** Ping Wang, Yeunjoo E. Song, Audrey Lynn, Kristy Miskimen, Alex Gulyayev, Michael B. Prough, Daniel A. Dorfsman, Renee A. Laux, Sarada L. Fuzzell, Sherri D. Hochstetler, Andrew F. Zaman, Larry D. Adams, Laura J. Caywood, Jason E. Clouse, Sharlene D. Herington, Patrice Whitehead, Yining Liu, Noel Moore, Paula Ogrocki, Alan J. Lerner, Anthony J. Griswold, Jeffery M. Vance, Michael L. Cuccaro, William K. Scott, Margaret A. Pericak-Vance, Jonathan L. Haines

## Abstract

**INTRODUCTION:** Alzheimer disease (AD) plasma biomarkers are noninvasive measures of the key amyloid beta (Aβ) and tau pathologies. Validation and generalization studies are needed to fully understand their potential for AD prediction and diagnosis in the elderly population.

**METHODS:** In 1,067 Amish individuals aged ≥ 65, we measured plasma Aβ and tau to assess their relationships with AD-related outcomes.

**RESULTS:** Among Amish individuals with AD, plasma p-tau181 was significantly higher (*p* = 0.04), and plasma Aβ42/p-tau181 ratio was significantly lower (*p* = 0.01) than cognitively normal individuals. The association of AD with elevated p-tau181 was driven by APOE ε4 carriers (OR = 6.02, *p* < 0.001). Cluster analysis identified two subgroups defined by differing Aβ and tau levels, with the high-risk cluster having more APOE ε4 carriers (*p* < 0.001).

**DISCUSSION:** Plasma biomarkers, particularly p-tau181, Aβ42/Aβ40, and Aβ42/p- tau181 ratio, are promising surrogate biomarkers for AD-related pathology and clinical outcomes in the Amish.

## 1. Background

Alzheimer disease (AD) is the leading cause of dementia, affecting approximately 6.9 million Americans aged 65 and older.^1^ The primary pathological hallmarks of AD include aggregates of amyloid beta (Aβ) plaques and tau tangles.^2,3^ Biomarkers reflecting these biological changes are critical to diagnosing AD and monitoring disease progression.^4^ Recent advances, including the approval of new AD therapeutics primarily targeting Aβ plaques,^5,6^ have established biomarkers as essential screening tools to identify eligible individuals for novel treatments. Although brain Aβ and tau pathologies can be reliably quantified *in vivo* using neuroimaging and cerebrospinal fluid (CSF) measures, the high cost, low accessibility, and invasiveness of these tests hinder population-level application. Plasma biomarkers, in contrast, offer a promising solution by mitigating these limitations. They can serve as accessible blood-based surrogates for AD pathologies and facilitate the identification of individuals at the earliest disease stage when the treatments are most likely to be successful.

The Aβ42/Aβ40 ratio and phosphorylated tau protein at epitope 181 (p-tau181) are promising plasma biomarkers for capturing abnormal Aβ and tau buildup. Plasma Aβ42/Aβ40 and p-tau181 have demonstrated good accuracy in differentiating between image-confirmed amyloid-positive and amyloid-negative individuals, as well as between individuals who are cognitively normal, have mild cognitive impairment (MCI), or are clinically diagnosed with AD and other forms of dementia.^7–10^ Circulating levels of these biomarkers are associated with neurodegeneration^11–18^ and the severity of dementia;^19,20^ they can also track disease progression and predict the onset of AD in individuals who were cognitively normal or had mild symptoms.^11,15,16,21–26^ However, most of these validation studies were conducted in clinical cohorts. A significant potential advantage of blood-based biomarkers is their ability to screen for AD pathological changes in at-risk populations at low burden and low cost. There is a great need to better understand the value of blood-based biomarkers in community-based elderly individuals.

We analyzed plasma biomarkers collected from the Amish communities in the Midwest (Ohio and Indiana), which present an ideal population for genetic studies of AD due to their unique genetic and cultural background.^27^ The Amish, with their tendency to marry those of the same faith, show less genetic diversity than the general population. Moreover, they share relatively uniform environmental exposures, including similar lifestyles, education, diets, and health behaviors. Previous surveys indicated a lower prevalence of cognitive impairment in the Amish compared to the general population, despite lower levels of formal education.^28–31^ However, AD plasma biomarkers have not been studied in the Amish. In this study, we focused on describing plasma Aβ42/Aβ40 and p-tau181 in relation to AD-related outcomes. Additional biomarkers, including the individual levels of plasma Aβ40, Aβ42, total tau (t-tau), and two Aβ/tau ratios (Aβ42/t-tau and Aβ42/p-tau181), were also investigated. Furthermore, we applied a clustering analysis using Aβ42/Aβ40 and p-tau181 to identify potential subgroups with unique biomarker profiles. This characterization and profiling of plasma biomarkers in the Amish will enhance our understanding of plasma biomarkers in the aging population.

## 2. Methods

### 2.1 Data Collection and Cognitive Testing

Data were collected from individuals enrolled in the Collaborative Amish Aging and Memory Project (CAAMP). CAAMP is a multi-center prospective study focusing on successful aging and dementia among the Amish population in Ohio and Indiana. The recruitment and ascertainment process for the CAAMP study has been previously described.^32,33^ A door-to-door interview was conducted to collect blood samples and data on demographics, medical history, neuropsychological function, and social determinants. We re-assessed individuals who were cognitively normal or had mild cognitive impairment at two-year intervals, and additional blood samples were obtained if possible. Genealogical records were available in the Anabaptist Genealogy Database to determine the degree of relatedness among Amish individuals.^34^ Genotype data were obtained from the Illumina Expanded Multi-Ethnic Genotyping Array (MEGAEX) with custom content or Illumina Global Screening Array (GSA). We confirmed the Apolipoprotein E (APOE) ε2/ε3/ε4 polymorphisms from the chip data and whole genome sequencing. All participants underwent a screening of cognitive function using the Modified Mini-Mental State Examination (3MS).^35^ The 3MS score ranges from 0 to 100, and individuals with a lower 3MS score are more likely to have cognitive deficits or early-stage dementia. A subset of Amish individuals underwent more in-depth assessments, including the CERAD Word List Learning Test,^36^ Verbal Fluency,^37^ Trail Making Test (A and B),^38^ Logical Memory,^39^ and Multilingual Naming Test.^40^

### 2.2 Clinical outcomes

Based on medical history and cognitive measurements, a clinical adjudication team consisting of neurologists and neuropsychologists classified participants as cognitively unimpaired (CU), mild cognitive impairment (MCI), cognitively impaired but not AD (CINAD), or cognitively impaired related to AD (AD). For our analyses, individuals who were MCI, CINAD, and AD were grouped together as cognitively impaired (CI). We defined two AD-related binary outcomes: 1) the AD outcome by comparing the subset of AD individuals to those who were CU, and 2) the cognitive preservation (CP) outcome by comparing CU individuals to CI individuals. For analyses involving CI, we conducted a sensitivity analysis by excluding individuals with MCI from this group.

### 2.3 Plasma biomarkers and quality control

Biomarker data were obtained from frozen plasma samples stored in 500ul aliquots at −80 degrees. Samples were randomly assigned on each plate and run in duplicate. Concentrations of plasma Aβ40, Aβ42, and t-tau were measured using the Simoa™ Neurology 3-Plex A or Neurology 4-Plex E assays (Quanterix), whilst p-tau181 was measured using the Simoa™ P-tau181 Advantage V2 assay. Results for all Aβ and tau biomarkers were above the detection limit of the assays. To ensure the quality of biomarkers used for analyses, we performed quality control (QC) at the individual biomarker level (Supplemental Figure 1). We first matched the biomarker samples to clinical data in the Amish. Samples without a consensus diagnosis or from non-Amish individuals were excluded. Some individuals had duplicate results from samples collected on the same date. In such cases, we excluded the set of results with a higher intra-assay coefficient of variation (CV). We then kept samples with a CV of less than 20%. Based on summary statistics and visualization, we evaluated samples for each biomarker on a case-by-case basis. The outlier for each biomarker was defined by the Median Absolute Deviation (MAD). This method is more robust in detecting outliers for skewed distributions due to its reliance on non-parametric measures of central tendency and variation.

### 2.4 Statistical analysis

From biomarkers that passed QC, we restricted our data to the most recently measured biomarkers for cross-sectional analysis. To compare differences in demographics, clinical features, and biomarkers between diagnostic groups (CU, MCI, CINAD, and AD), we used the Kruskal-Wallis test for continuous variables with non-parametric distributions. For categorical variables, we used either the Chi-square test or Fisher’s exact test. We applied the Bonferroni method to correct for multiple comparisons in the post-hoc analysis.

Relationships between plasma biomarkers were estimated using the Spearman’s rank correlation coefficient. Due to the skewed distributions of biomarker data, plasma biomarkers were log-transformed to better approximate normality and variance homogeneity. To study the association between each biomarker and AD, we applied logistic regression in individuals with AD and CU, adjusting for age at sample collection, age^2^, sex, presence/absence of APOE ε4 alleles, time from specimen collection to diagnosis, biomarker batches, and research center. To understand the predictive value of biomarkers for cognitive preservation (CP) in the CU individuals, we performed logistic regression in the CU and CI groups, adjusting for the covariates above but replacing APOE ε4 allele status with the presence/absence of APOE ε2. The interaction between biomarkers and APOE (ε4 or ε2) was included as an additional covariate in all multivariate analyses. We evaluated the performance of plasma biomarkers in classifying AD and CP using the receiver operating characteristic curve (ROC) analysis. The optimal cutoff value was determined by the Youden’s Index. Sensitivity, specificity, positive predictive value (PPV), negative predictive value (NPV), and areas under the curve (AUC) were reported. We also applied the 10-fold cross-validation to assess the model’s stability across different subsets of the data and ensure robust generalization.

To identify Amish subgroups, we applied the unsupervised K-medoids clustering approach using plasma Aβ42/Aβ40 and p-tau181. Potential confounding effects were addressed by regressing out the influences of age, sex, and research center using a quadratic regression model. Adjusted biomarkers were then inverse normalized for the clustering analysis. The distance matrix was calculated using the Euclidean distance. The K-medoids method is more robust in minimizing the distance between points in the same cluster. The primary analysis set the number of clusters at two to test if plasma Aβ42/Aβ40 and p-tau181 may split the Amish population into two groups. We also checked the optimal number of clusters derived from the elbow method and Silhouette score as a secondary analysis. Good clustering is indicated by the elbow point of the scree plot and a high average silhouette width. To interpret the cluster profiles, we explored the differences in demographics and clinical features between clusters. We further evaluated the relationships between clusters and cognitive function after controlling for diagnosis, sex, age, age^2^, research center, and APOE ε4 allele status. All statistical analyses were conducted in R (version 1.4.1717, https://www.r-project.org).

## 3. Results

### 3.1 Study population

After quality control, the cross-sectional data set consisted of 1,067 individuals for biomarker analysis. The average age was 81.2 ± 6.2 years. 50.4% of the individuals were from the Ohio research center, 40.9% were males, and 74.1% had eight years of education. The characteristics of these individuals by diagnostic groups (i.e., CU, MCI, CINAD, or AD) are presented in Table 1. Compared to CU individuals, individuals with AD or MCI were significantly older (*P_Bonferroni_* < 0.001). 268 individuals carried at least one APOE ε4 allele, and comparing across diagnostic groups, 46.4% of the AD individuals were APOE ε4 carriers, which is significantly higher than the percentage of carriers in the CU (*P_Bonferroni_* < 0.001) or MCI group (*P_Bonferroni_* = 0.003).

### 3.2 Univariate analysis of plasma biomarkers

Detailed results of plasma biomarkers by consensus diagnosis are included in Supplemental Table 1. No significant differences were found in plasma amyloid biomarkers (i.e., Aβ40, Aβ42, Aβ42/Aβ40) between diagnostic groups. As shown in Figure 1, after adjusting for multiple comparisons, individuals in the AD group had a significantly higher level of plasma p-tau181 than CU individuals (2.1 ± 0.9 pg/mL vs. 1.9 ± 0.8 pg/mL, *P_Bonferroni_* = 0.04). For the ratio of plasma Aβ42/p-tau181, individuals with AD had significantly lower Aβ42/p-tau181 (3.6 ± 2.6) compared to individuals with CINAD (5.5 ± 22.9, *P_Bonferroni_* = 0.02) and CU (4.8 ± 2.9, *P_Bonferroni_* = 0.01). Plasma t-tau and Aβ42/t-tau ratio did not differ significantly between groups.

**Figure 1.**
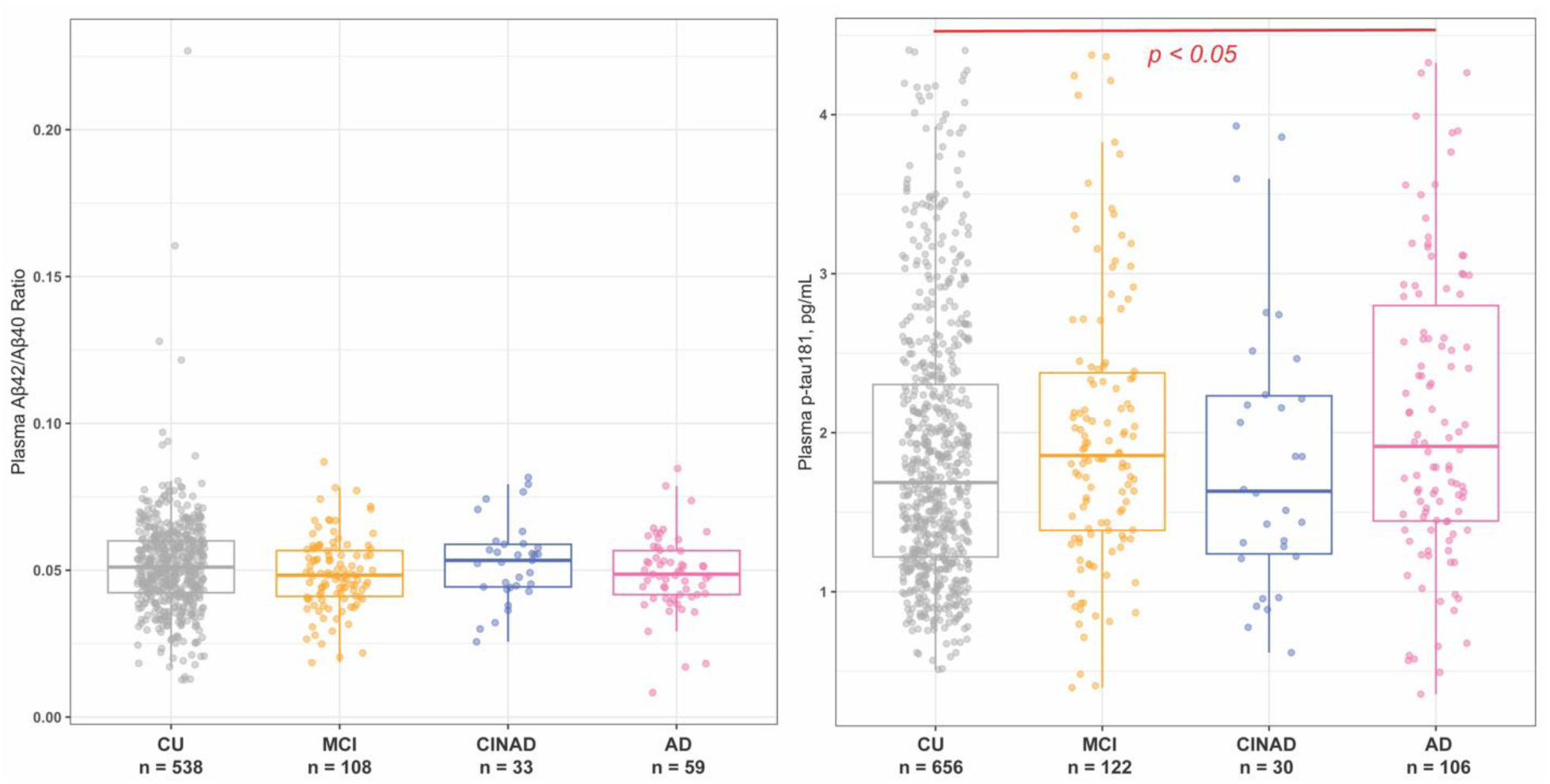
Plasma p-tau181, but not Aβ42/Aβ40 ratio, was significantly higher in Amish individuals with AD than CU individuals. Abbreviations: CU, cognitively unimpaired; MCI, mild cognitive impairment; AD, Alzheimer disease; CINAD, cognitively impaired but not AD.

**Table 1.** Participant characteristics by consensus diagnosis.

|  | CU | MCI | CINAD | AD | *p*<sup>*</sup> |
| --- | --- | --- | --- | --- | --- |
| N | 754 | 147 | 40 | 126 |  |
| Ohio Center, n (%) | 413 (54.8) | 61 (41.5) | 12 (30.0) | 66 (52.4) | 0.001<sup>†</sup> |
| Age, Mean (SD) | 80.3 (6.3) | 83.1 (4.9) | 82.0 (7.9) | 83.5 (5.6) | < 0.001<sup>‡</sup> |
| Male, n (%) | 289 (38.3) | 68 (46.3) | 22 (55.0) | 49 (38.9) | 0.07 |
| APOE ε4 Carrier, n (%) | 164 (22.2) | 36 (25.2) | 10 (25.6) | 58 (46.4) | < 0.001<sup>§</sup> |
| APOE Genotype, n (%) |  |  |  |  | < 0.001<sup>¶</sup> |
| ε2/ε2 | 0 ( 0.0) | 1 ( 0.7) | 0 ( 0.0) | 0 ( 0.0) |  |
| ε2/ε3 | 75 (10.2) | 7 ( 4.9) | 5 (12.8) | 10 ( 8.0) |  |
| ε2/ε4 | 11 ( 1.5) | 2 ( 1.4) | 3 ( 7.7) | 3 ( 2.4) |  |
| ε3/ε3 | 499 (67.6) | 99 (69.2) | 24 (61.5) | 57 (45.6) |  |
| ε3/ε4 | 144 (19.5) | 30 (21.0) | 7 (17.9) | 47 (37.6) |  |
| ε4/ε4 | 9 ( 1.2) | 4 ( 2.8) | 0 ( 0.0) | 8 ( 6.4) |  |
| Education, n (%) |  |  |  |  | 0.08<sup>¶</sup> |
| < 8 years | 56 ( 7.4) | 6 ( 4.1) | 3 ( 7.5) | 16 (12.7) |  |
| 8 years | 546 (72.4) | 114 (77.6) | 30 (75.0) | 95 (75.4) |  |
| > 9 years | 111 (14.7) | 25 (17.0) | 5 (12.5) | 10 ( 7.9) |  |

\* *p*-values were obtained from the Kruskal-Wallis, Chi-square or Fisher's exact tests.
† After adjusting for multiple comparisons, the percentage of Ohio residents was significantly higher in the CU group than the MCI or CINAD group.
‡ After adjusting for multiple comparisons, the CU individuals were significantly younger than individuals with MCI or AD.
§ After adjusting for multiple comparisons, the percentage of APOE ε4 carriers was significantly higher in the AD group than the CU or MCI group.
¶ *p*-value was obtained from the Fisher's exact test.
Abbreviations: SD, standard deviation; CU, cognitively unimpaired; MCI, mild cognitive impairment; AD, Alzheimer disease; CINAD, cognitively impaired but not AD; APOE, apolipoprotein E.

Among all individuals, the correlations between Aβ biomarkers were significant (*p* < 0.05), and plasma Aβ40 and Aβ42 had the strongest positive correlation (ρ = 0.83) (Supplemental Figure 2). t-tau and p-tau181 also were significantly correlated (ρ = 0.24, p < 0.05). Between plasma Aβ and tau biomarkers, t-tau had significant correlations with Aβ40 (ρ = 0.29, *p* < 0.05) and Aβ42 (ρ = 0.25, *p* < 0.05), but the correlations between p-tau181 and Aβ biomarkers were weak. Moderate to strong correlations (ρ ranging from 0.32 - 0.49) were found between the different ratios. The correlation matrix in CU individuals had a similar pattern. However, among individuals with AD, we found less significant correlations by *p*-values.

### 3.3 Multivariate analysis of plasma biomarkers and AD-related outcomes

We examined the relationships between each plasma biomarker and AD-related outcomes after adjusting for covariates, including age, age^2^, sex, the presence of APOE (ε2 or ε4) alleles, time from specimen collection to diagnosis, biomarker batches, research center, and the interaction between biomarkers and APOE (Supplemental Table 2). No statistically significant associations were found between plasma biomarkers and AD in the multivariate analyses (Figure 2A). Amish individuals with AD tended to have lower Aβ biomarkers in the plasma, including Aβ42/Aβ40, Aβ40, and Aβ42. The ratios of Aβ42/t-tau and Aβ42/p-tau181 showed similar trends of associations. In contrast, AD individuals were more likely to have a higher plasma t-tau. Interestingly, the association between p-tau181 and AD (OR = 0.77, 95% CI: 0.39 - 1.51, *p* = 0.44) was modulated by the interaction between p-tau181 and APOE ε4 (OR = 8.55, 95% CI: 2.69 - 28.92, *p* = 0.0004). After stratifying by APOE ε4 (Supplemental Table 3), p-tau181 was not significantly associated with AD among non-carriers (OR = 0.75, 95% CI: 0.38 - 1.94, *p* = 0.41). Nonetheless, among APOE ε4 carriers AD was strongly associated with increased levels of p-tau181 (OR = 6.02, 95% CI: 2.33 - 17.11, *p* < 0.001). As expected, for the cognitive preservation outcome (Figure 2B), plasma biomarkers essentially showed the opposite direction of associations compared to the results in the AD outcome. The strongest relationship was found between plasma Aβ42/Aβ40 and CP (OR = 1.81, 95% CI: 1.01 - 3.23, *p* = 0.05), which was no longer statistically significant in the sensitivity analysis excluding MCI from the CI group (Figure 2C). Notably, individuals with CP were significantly more likely to have decreased p- tau181 (OR = 0.58, 95% CI: 0.35 - 0.96, *p* = 0.04) and increased Aβ42/p-tau181 ratio (OR = 1.80, 95% CI: 1.18 - 2.76, *p* = 0.01) in the sensitivity analysis.

**Figure 2.**
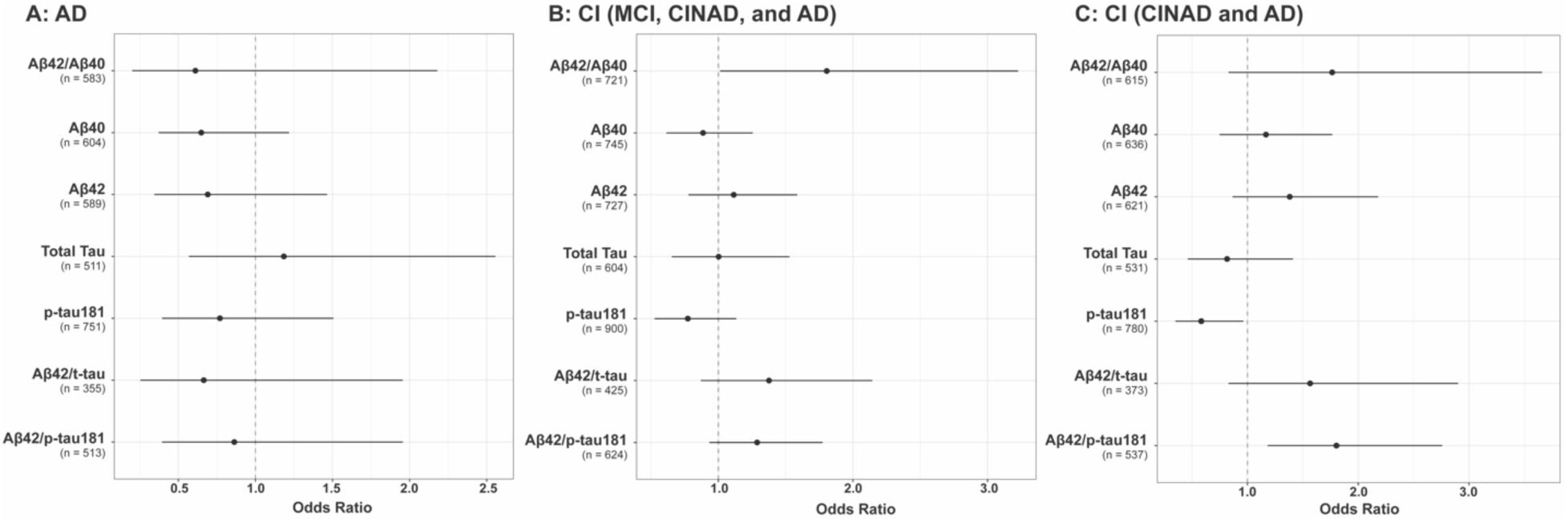
Forest plots illustrating the associations between plasma biomarkers and three clinical outcomes: plasma p-tau181 and the Aβ42/p-tau181 ratio were significantly associated with cognitive preservation when comparing CU individuals to those with CI (including CINAD and AD). A) Multivariate regression models were applied among AD and CU individuals. B) Multivariate regression models were applied among individuals with CU and CI (MCI, CINAD, and AD). C) Multivariate regression models were applied among individuals with CU and CI (CINAD and AD). Abbreviations: AD, Alzheimer disease; CP, cognitive preservation; CU, cognitively unimpaired; MCI, mild cognitive impairment; CINAD, cognitively impaired but not AD.

To evaluate the performance of plasma biomarkers in distinguishing AD from CU, we compared each model to a reference model (Supplemental Table 4). This reference model, adjusted for covariates of age, age^2^, sex, research center, and time from sample collection to diagnosis, yielded an AUC of 0.64. While the PPV was high, the sensitivity, specificity, and NPV values were low. As expected, incorporating APOE ε4 into the reference model generated a higher AUC and sensitivity. Furthermore, the addition of individual biomarkers generally improved the model’s discriminative performance for AD, except for t-tau. The best model in predicting AD was the one using all biomarkers or Aβ42/p-tau (AUC = 0.83). For the cognitive preservation outcome, adding individual biomarkers to the reference model did not meaningfully enhance the model’s performance. The most useful predictive model for CP was when APOE ε2 and all biomarkers were included (AUC = 0.71), followed by the one incorporating APOE ε2 and Aβ42/t-tau (AUC = 0.70). In the sensitivity analysis excluding MCI, models with APOE ε2 and individual biomarkers overall worked better with higher AUCs than analyses including MCI, except for t-tau (Supplemental Table 5). The predictive model for CP performed the best when APOE ε2 and all biomarkers (or Aβ42/pTau) were included (AUC = 0.77). In the 10-fold cross-validation of these models, the average performance matrices for each biomarker in classifying AD or CP were similar.

### 3.4 Clustering results using plasma Aβ42/Aβ40 and p-tau181

In the primary clustering analysis using a pre-fixed number of clusters (k = 2), Amish individuals can be divided into two subgroups based on plasma Aβ42/Aβ40 and p- tau181 (Figure 3). Cluster 1 (n = 382) showed a low-risk profile for AD characterized by elevated levels of plasma Aβ42/Aβ40 (0.06 ± 0.02) and reduced levels of p-tau181 (1.71 ± 0.63 pg/mL). In contrast, Cluster 2 (n = 294) presented a vastly different pattern with a higher burden of Aβ (decreased Aβ42/Aβ40) and tau (increased p-tau181), indicating a high-risk profile for AD. Notably, Aβ42 was significantly lower in Cluster 2 than Cluster 1 (7.17 ± 3.46 vs. 9.23 ± 3.64, *p* < 0.001). The concentration of plasma Aβ40 and t-tau, however, did not show significant differences between the two clusters.

**Figure 3.**
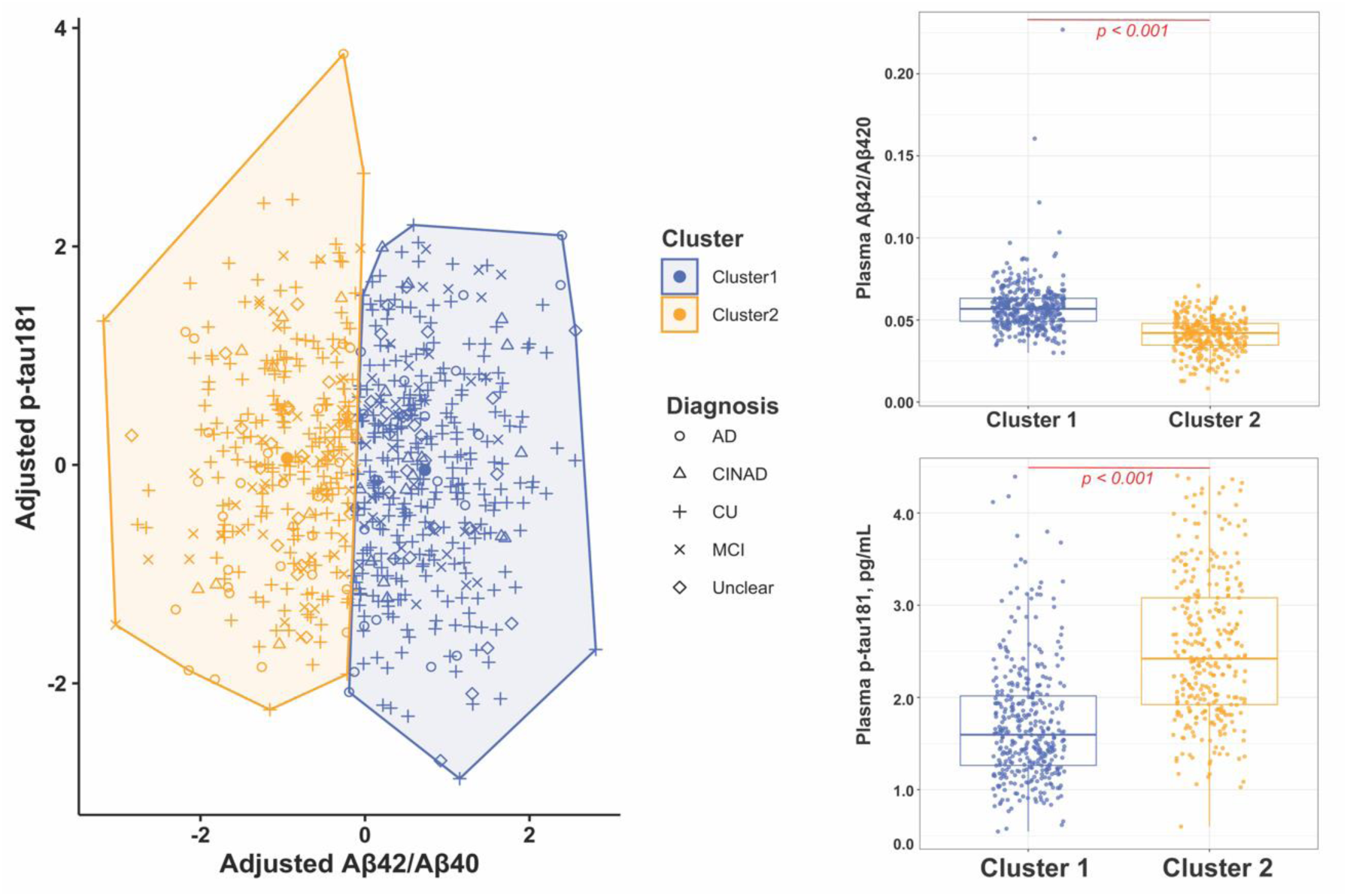
Clustering analysis based on plasma Aβ42/Aβ40 and p-tau181 levels categorized Amish individuals into two distinct groups (k = 2) with low-risk and high-risk profiles for AD. Abbreviations: CU, cognitively unimpaired; MCI, mild cognitive impairment; CINAD, cognitively impaired but not AD; AD, Alzheimer disease.

The two clusters did not differ significantly by age, sex, research center, education, and history of smoking (Table 2). However, Cluster 2 contained significantly more APOE ε4 carriers (*p* < 0.001) and individuals with AD (*p* = 0.04) than Cluster 1. Upon further examination (Supplemental Table 6), we found no significant differences between the two clusters in the self-reported medical history of neurological disorders or cardiovascular risk factors, such as Parkinson’s disease, multiple sclerosis, epilepsy, heart disease, diabetes, and hypertension. Cluster 1, however, had more individuals reporting a history of elevated lipids/cholesterol than Cluster 2 (*p* = 0.02). Although not significant, individuals with AD had a higher chance of being in Cluster 2 (OR = 1.55, 95% CI: 0.80 - 3.04) after controlling for age, age^2^, sex, research center, and APOE (Supplemental Table 7). Meanwhile, individuals with cognitive preservation were less likely to appear in Cluster 2, especially when MCIs were excluded from the analysis (OR = 0.60, 95% CI: 0.41 - 0.88, *p* = 0.01). No significant associations were found between the two clusters when cognitive test results (e.g., 3MS, verbal fluency, working memory, logical memory, MINT) were examined individually.

**Table 2.** Characteristics of pre-fixed number of clusters (k = 2) using plasma Aβ42/Aβ40 and p-tau181.

|  | Cluster 1 (n = 382) | Cluster 2 (n = 294) | *p* |
| --- | --- | --- | --- |
| Core Biomarkers, mean (SD) |  |  |  |
| Aβ42/Aβ40 | 0.06 (0.02) | 0.04 (0.01) | <0.001 |
| Aβ40 | 166.29 (66.75) | 176.21 (72.58) | 0.17 |
| Aβ42 | 9.23 (3.64) | 7.17 (3.46) | <0.001 |
| t-tau | 1.81 (0.72) | 1.86 (0.67) | 0.31 |
| p-tau181 | 1.71 (0.63) | 2.52 (0.79) | <0.001 |
| Demographic and Clinical Characteristics |  |  |  |
| Age, mean (SD) | 81.69 (5.96) | 81.97 (5.52) | 0.53 |
| Male, n(%) | 146 (38.2) | 120 (40.8) | 0.55 |
| Center (Ohio), n (%) | 132 (34.6) | 101 (34.4) | 0.99 |
| Education, n (%) |  |  | 0.33 |
| < 8 years | 24 ( 6.3) | 18 ( 6.1) |  |
| 8 years | 273 (71.5) | 207 (70.4) |  |
| 9 - 12 years | 61 (16.0) | 58 (19.7) |  |
| Smoking, n (%) |  |  | 0.44 |
| Never | 252 (66.0) | 204 (69.4) |  |
| <100 | 50 (13.1) | 31 (10.5) |  |
| >100 | 58 (15.2) | 48 (16.3) |  |
| APOE ε2 Carrier, n (%) | 45 (11.8) | 29 ( 9.9) | 0.39 |
| APOE ε4 Carrier, n (%) | 70 (18.3) | 103 (35.0) | <0.001 |
| APOE Genotype, n (%) |  |  | <0.001 |
| ε2/ε3 | 40 (10.5) | 21 ( 7.1) | |
| ε2/ε4 | 5 ( 1.3) | 8 ( 2.7) | |
| ε3/ε3 | 264 (69.1) | 167 (56.8) | |
| ε3/ε4 | 64 (16.8) | 86 (29.3) | |
| ε4/ε4 | 1 ( 0.3) | 9 ( 3.1) | |
| Diagnosis, n (%) |  |  | 0.04 |
| CU | 279 (73.0) | 191 (65.0) |  |
| MCI | 40 (10.5) | 48 (16.3) |  |
| CINAD | 16 ( 4.2) | 9 ( 3.1) |  |
| AD | 21 ( 5.5) | 27 ( 9.2) |  |
| Unclear | 26 ( 6.8) | 19 ( 6.5) |  |

Abbreviations: SD, standard deviation; APOE, apolipoprotein E; CU, cognitively unimpaired; MCI, mild cognitive impairment; CINAD, cognitively impaired but not AD; AD, Alzheimer disease.

In the secondary clustering analysis, the elbow and silhouette methods suggested an optimal number of three clusters in the Amish (Supplemental Figure 3). Between k = 2 and k = 3, the clustering results revealed a clearer distinction between Aβ and tau profiles, particularly in the distribution of moderate and low levels across individuals. At k = 3, the original Cluster 1 from the primary clustering analysis was evenly divided into two distinct subclusters: one cluster (n = 192) exhibited a profile of low Aβ risk, characterized by the highest Aβ42/Aβ40 ratio (0.06 ± 0.02) and Aβ42 level (10.01 ± 3.95 pg/mL), but high Tau risk with a high p-tau181 level (2.20 ± 0.71pg/mL); the other cluster (n = 190) showed a moderate Aβ risk profile with slightly lower Aβ42/Aβ40 ratio of 0.05 ± 0.01 and Aβ42 level of 7.87 ± 3.16 pg/mL, alongside a low Tau risk evidenced by the lowest p-tau181 level of 1.41 ± 0.41 pg/mL. Individuals who were previously grouped together with low levels of Aβ and Tau exhibited greater differences in tau levels, resulting in their placement in distinct clusters. Similarly, Cluster 2 from the primary clustering analysis was distributed across all three clusters at K=3: the one characterized by a profile of low Aβ risk and high Tau risk (n = 34), another by a moderate Aβ risk and low Tau risk profile (n = 49), as described above. The remaining individuals (n = 211) retained a profile of high Aβ risk and high Tau risk, characterized by the lowest level of Aβ42 (7.05 ± 3.32 pg/mL) and Aβ42/Aβ40 ratio (0.04 ± 0.01), along with the highest level of p-tau181 (2.67 ± 0.72 pg/mL). This more refined clustering provides a nuanced differentiation between individuals with low Aβ but high tau levels and those with moderate Aβ and low tau levels.

As shown in Supplemental Table 8, Cluster 2 had a significantly lower level of p-tau181 than Cluster 1 and 3. From Cluster 1 to Cluster 3, there was a significantly increasing burden of amyloid suggested by decreasing levels of Aβ42/Aβ40 and Aβ42. In Cluster 3, 40.4% of individuals were APOE ε4 carriers, a proportion significantly higher compared to the other two clusters (*p* < 0.001). None of the medical histories differed significantly between clusters (Supplemental Table 9). There were no significant associations between AD-related outcomes and clusters after adjusting for age, age^2^, sex, research center, and the presence of APOE alleles (ε4 for AD and ε2 for CP). In models adjusting for the above covariates and diagnosis, Cluster 3 was associated with a prolonged completion of Trails Making B (beta = 15.8 seconds, 95% CI: 0.53 - 31.09, *p* = 0.04). However, this association did not survive multiple testing (Supplemental Table 10).

## 4. Discussion

In this cross-sectional study, we describe the characteristics of AD core plasma biomarkers in the Amish, including Aβ40, Aβ42, Aβ42/Aβ40, t-tau, p-tau181, Aβ42/t-tau, and Aβ42/p-tau181. Compared to cognitively unimpaired individuals, those with AD had significantly higher p-tau181 and lower Aβ42/p-tau181 ratio. We explored the value of these biomarkers by analyzing their associations with both AD and cognitive preservation outcomes, in which plasma p-tau181 emerged as the most promising biomarker, demonstrating a statistically significant association with AD among APOE ε4 carriers. Furthermore, p-tau181 and Aβ42/p-tau181 ratio were also associated with the CP outcome among individuals with CU, AD, and CINAD. Through the k-medoids approach, we first tested an a priori hypothesis that there were two Amish subgroups with unique Aβ and tau profiles using plasma Aβ42/Aβ40 and p-tau181. This clustering was refined when using three clusters. To our knowledge, this is the first study to summarize AD-related plasma biomarkers for amyloid-beta and tau pathologies in the Amish. Founder populations like the Amish, characterized by unique genetic homogeneity, reduced environmental variability, and a lower prevalence of common confounding factors, offer a valuable opportunity to explore robust associations between biomarkers and AD risk and resilience.

In plasma, neither Aβ40, Aβ42, or Aβ42/Aβ40 differed significantly between diagnostic groups. However, as expected, Aβ42/Aβ40 emerges as a promising biomarker in associating with AD, surpassing the individual Aβ40 or Aβ42. The ratio can adjust for between-individual differences in total Aβ production, therefore, having higher concordance with brain amyloidosis.^41^ Numerous studies have shown that plasma Aβ42/Aβ40 has a high correspondence with amyloid PET positivity,^42,43^ and a lower Aβ42/Aβ40 ratio was significantly associated with the onset of AD and long-term cognitive deterioration.^44,45^ Although not statistically significant, we observed inverse relationships between Aβ biomarkers and AD after adjusting for covariates, aligning with the previously reported trends. Notably, elevated plasma Aβ42/Aβ40 was associated with cognitive preservation in the Amish (OR = 1.81, *p* = 0.05), suggesting its potential use to differentiate normal aging and cognitive impairment.

We evaluated t-tau and p-tau181 to reflect neuronal damage and subsequent drainage of tau from the brain parenchyma to blood. We found that plasma p-tau181 was elevated in the Amish individuals with AD (*p* = 0.04). Furthermore, we found a significant interaction between AD and log-transformed p-tau181 and APOE ε4 in the multivariate regression model (*p* < 0.001). AD was associated with a higher level of p- tau181 only in APOE ε4 carriers (OR = 6.02, *p* < 0.001). The moderating effect of APOE ε4 on the association of AD and plasma p-tau181 was in line with previous studies, wherein plasma p-tau181 levels were strongly associated with AD,^46^ neurodegeneration,^46,47^ memory deficits,^47^ and cognitive decline,^48^ almost exclusively in the APOE ε4 carriers. The mechanism by which APOE interacts with plasma p-tau181 is not clear. APOE ε4 may exacerbate tau pathology, possibly by affecting tau aggregation, phosphorylation, or tau-mediated neurodegeneration.^49,50^ Overexpression of APOE ε4 in astrocytes can increase the phosphorylation and aggregation of tau within neurons.^51^ APOE is also recognized for its role in modulating the blood-brain barrier (BBB); ε4 allele damages the integrity of the BBB, which may facilitate the escape of p-tau181 into CSF and plasma.^52^ Notably, the correlation of APOE ε4 and tau is likely independent of Aβ pathology. Shi et al. showed that tau pathogenesis and tau-mediated neurodegeneration were more severe in mouse models transfected with the human ApoE4 gene without influence from Aβ.^53^ Thus, regardless of the disease staging, APOE ɛ4 carriers with elevated p-tau181 may be more susceptible to the onset and progression of AD. Despite a significant positive correlation between t-tau and p- tau181 (ρ = 0.24, *p* < 0.05), t-tau did not vary much between diagnostic groups and had no associations with AD or cognitive preservation in the Amish. Additionally, regression models integrating t-tau performed the poorest in identifying AD or CP according to AUCs. Plasma t-tau is not specific to AD. Elevated t-tau levels can be observed in various neurodegenerative diseases and neurological conditions leading to neuronal damage.^54^ Plasma t-tau is also age-dependent.^55,56^ Individuals enrolled in this study were primarily older adults; the large overlap between normal aging and dementia may play a role in the non-significant findings for plasma t-tau in the Amish.

Since AD is characterized by multiple pathological changes, a combination of plasma biomarkers reflecting co-occurring pathology may be more powerful in classifying people at risk for AD diagnostically. In our models distinguishing between AD and CU, the model incorporating all biomarkers (i.e., Aβ40, Aβ42, Aβ42/Aβ40, t-tau, and p- tau181) outperformed the models using individual biomarkers alone based on AUCs. For the cognitive preservation outcome, we also observed improved AUCs and sensitivity values using all biomarkers. Interestingly, we found that plasma Aβ42/p- tau181 was a superior measure in classifying AD than individual biomarkers as well. Aβ42/p-tau181 was significantly decreased in the Amish individuals with AD (*p* = 0.01), suggesting concomitant AD pathology. Elevated Aβ42/p-tau181 ratio was associated with higher odds of cognitive preservation among CU and CI (i.e., CINAD and AD) individuals. Moreover, Aβ42/p-tau181 had similar discriminative capability for AD and CP compared to the models using all biomarkers, corroborating findings from previous studies that Aβ42/p-tau181 added discriminatory power over solitary Aβ42, t-tau and p- tau181 in separating AD from controls and other dementias.^57^ ^58^ Our results provide additional evidence supporting the use of plasma Aβ42/p-tau181 to assess AD and cognitive preservation in the aging population.

Alternatively, we integrated plasma Aβ42/Aβ40 and p-tau181 in the clustering analysis to detect the heterogeneity in biomarker profiles regardless of diagnoses. Common methods for determining the optimal number of clusters, such as the elbow method and silhouette analysis, rely heavily on summary statistics (e.g., within-cluster sum of squares, average silhouette score). Such metrics may not effectively capture the underlying clustering structure, especially in small datasets.^59^ Additionally, small sample sizes amplify the risk of spurious results due to noise or outliers. Given the limited number of individuals in the Amish with both plasma Aβ42/Aβ40 and p-tau181 measurements, we chose k=2 as the primary clustering solution. This choice simplifies interpretation and provides clearer insights into the variability of biomarker profiles within the Amish population. Furthermore, using two clusters ensures larger cluster sizes, which enhances statistical power for subsequent analyses. The primary clustering analysis divided Amish individuals into two subgroups, with one group displaying a high- risk profile for AD-related pathology, evidenced by the decreased levels of Aβ42/Aβ40 and Aβ42 in combination with increased p-tau181. The other group featured a low-risk profile with relatively higher levels of Aβ42/Aβ40 and Aβ42, and a lower level of p- tau181. When separating the Amish into three clusters as indicated by the elbow plot and Silhouette width, individuals were further separated based on decremental levels of Aβ42/Aβ40 and Aβ42. Either way, the subgroup with the highest Aβ and tau burden consisted of a significantly higher proportion of APOE ε4 carriers (*p* < 0.001), and this group also tended to have more individuals with AD. The clusters did not differ significantly in age, sex, education, smoking, and medical history. While assessing the association between method-derived clusters and the Trails Making Test B, Cluster 3 with a high-risk profile for Aβ and tau had worse performance than individuals in Cluster 1 with a relatively lower risk for Aβ but still high risk for tau. Although this relationship did not survive the multiple comparisons, our data might suggest an independent association between Aβ and executive function. Previous studies in preclinical AD have shown the relationship between Aβ accumulation and executive function preceding memory impairment,^60,61^ implying the importance of early screening using both executive and memory measurements.

A few limitations should be acknowledged in this work. First, the Amish population is relatively small and is not representative of the general aging population with a European ancestry. Second, the clinical diagnosis was derived mainly from the medical history and cognitive measurements as the Amish do not have imaging or CSF data available, there might be a misclassification of diagnosis, especially for individuals with MCI or subtle impairments. Third, the plasma biomarker levels are only a proxy for AD pathologies in the brain. However, reliable assays are now available to measure extremely low levels of brain-derived proteins in blood samples^62,63^ and alterations in plasma biomarkers are in accordance with the changes in CSF.^64^ We applied rigorous quality control at the research center and biomarker level, but other pre-analytical factors should be taken into consideration while harmonizing biomarker data from different centers.^65^ Lastly, the cross-sectional analysis only provided a snapshot of the associations between plasma biomarkers and AD-related outcomes; longitudinal data to understand the changes in plasma biomarkers over time are critical to monitor the development of AD. Despite these limitations, this study is novel in characterizing and profiling core AD plasma biomarkers in the elderly Amish population. Due to the reduced genetic diversity and relatively homogeneous environment, Amish may be an ideal population to study the genetics of plasma biomarkers to advance the understanding of AD pathophysiology. Examinations of genetic variants influencing plasma biomarkers in the Amish are underway.

## 5. Conclusions

In summary, plasma biomarkers, especially p-tau181, Aβ42/Aβ40, and Aβ42/p-tau181, are promising non-invasive indicators to distinguish Amish individuals with AD or cognitive impairments from those who are cognitively unimpaired. Additionally, we demonstrate that biomarker-guided clustering using Aβ42/Aβ40 and p-tau181 can identify individuals at various levels of risk for AD. These findings are important to screen for biomarker evidence of AD in older adults and potentially form homogenous subgroups that share similar disease mechanisms and treatment responses.

Longitudinal studies are needed to validate the Aβ, tau, and neurodegeneration biomarkers in early detection and disease monitoring in the Amish and older population.

## Data Availability

All data produced in the present study are available upon reasonable request to the authors.

## Acknowledgements

This research was supported under the National institutes of Health and National Institute on Aging (grants AG019085, AG019726, and AG058066).

## Consent Statement

All study procedures were approved by the Institutional Review Board at the Case Western Reserve University and University of Miami. Written informed consent was obtained from the participant or legal guardian. For the sake of confidentiality, clinical and genetic data were de-identified according to the study protocol.

---

**Supplemental Figure 1.**
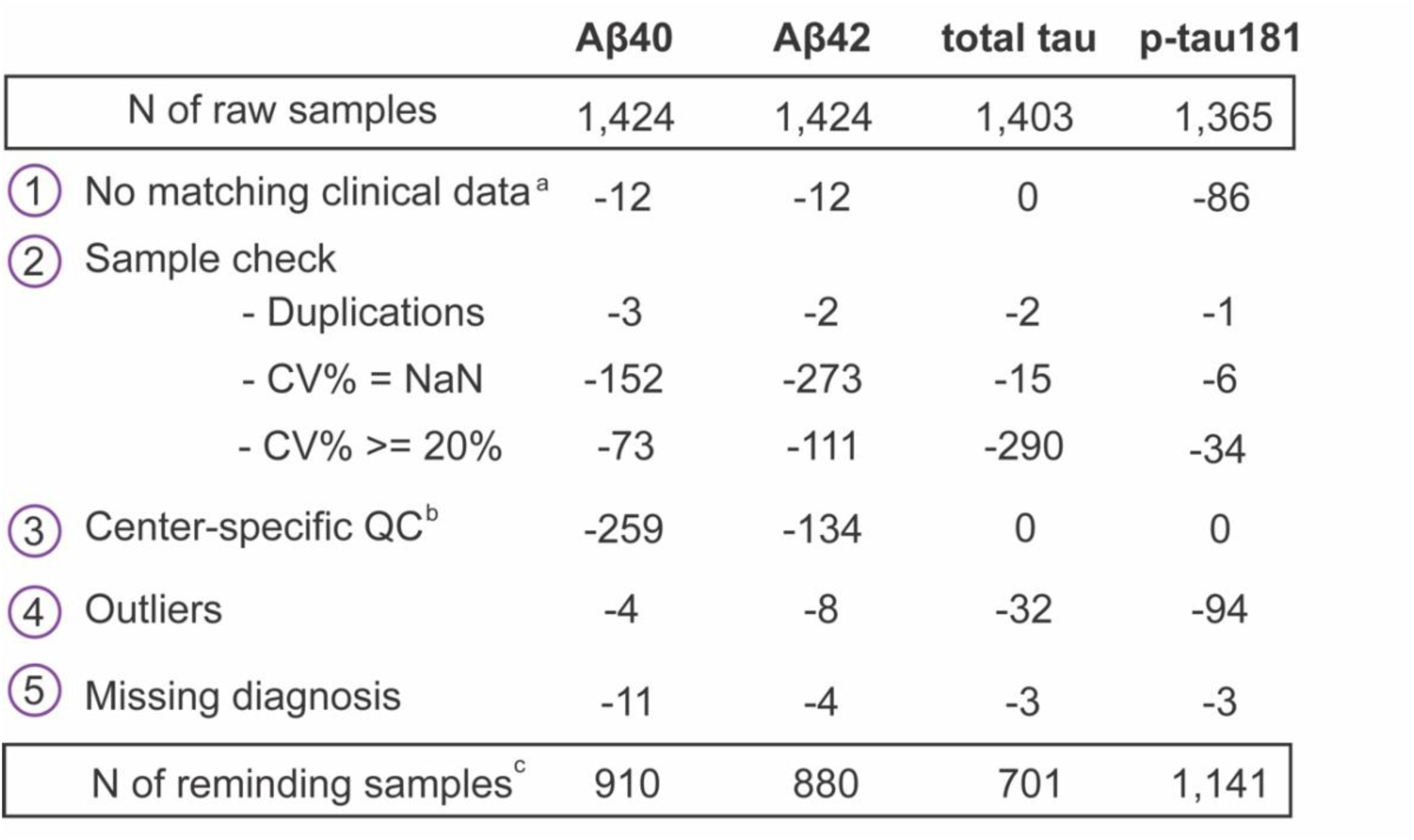
Biomarker-level quality control. a. Samples had no matching clinical data (i.e., no consensus diagnosis or not Amish). b. Aβ40 and Aβ42 samples before 2014 from Ohio were excluded. c. Samples passed QC include one-time and repeated measurements on the same individual. Abbreviations: QC, quality control; CV, Coefficient of variation percentages.

**Supplemental Figure 2.**
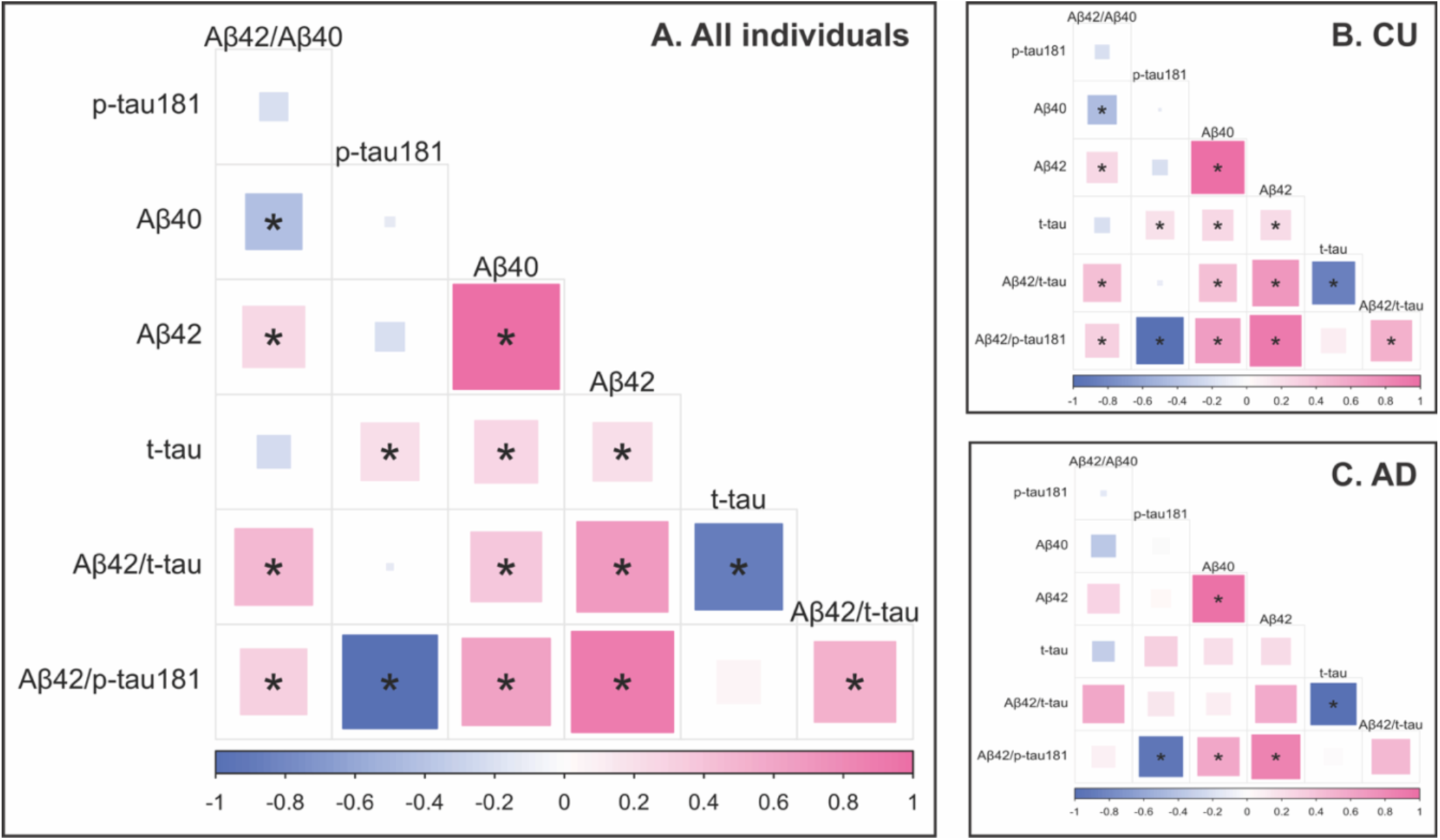
Correlations between the most recently measured plasma biomarkers. Spearman’s Rank correlation was performed between biomarker pairs among A) all individuals with a clear diagnosis, B) CU individuals, and C) AD individuals. Pink shading indicates positive correlations, and blue shading indicates negative correlations. The size of the squares indicates varying levels of significance. The darker color indicates a stronger correlation. A statistically significant correlation (*p* < 0.001) after Bonferroni correction is shown by asterisk symbols. Abbreviations: CU, cognitively unimpaired; AD, Alzheimer disease.

**Supplemental Figure 3.**
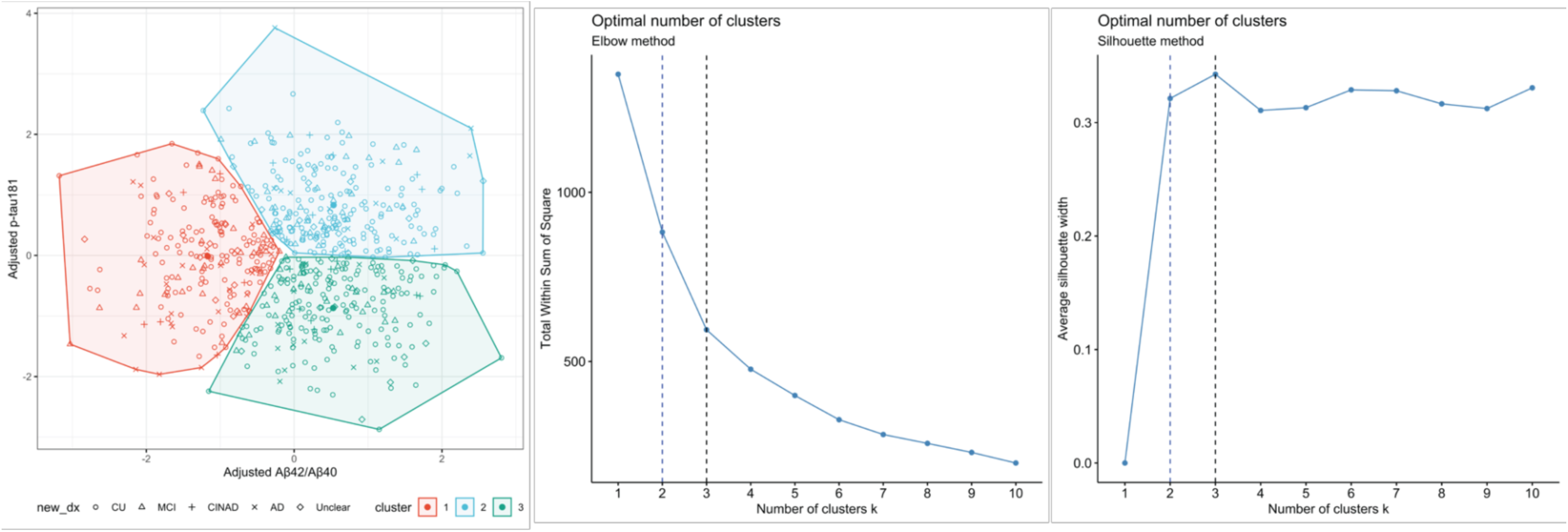
Determination of the optimal number of clusters using the elbow and silhouette methods. Note: The optimal number of clusters was assessed by the elbow method (Middle) and silhouette width (Right). The curve in the elbow plot started to level off after k = 3 by a visual examination. The highest silhouette width was reached when k = 3, followed by k = 2.

**Supplemental Table 1.** Comparison of plasma beta-amyloid and tau biomarkers between diagnostic groups.

| Unit: pg/mL | CU |  | MCI |  | CINAD |  | AD |  | *p* * |
| --- | --- | --- | --- | --- | --- | --- | --- | --- | --- |
|  | N | Mean (SD) | N | Mean (SD) | N | Mean (SD) | N | Mean (SD) |  |
| Aβ42/Aβ40 | 538 | 0.052 (0.017) | 108 | 0.049 (0.012) | 33 | 0.053 (0.014) | 59 | 0.049 (0.013) | 0.15 |
| p-tau181 | 656 | 1.9 (0.8) | 122 | 2.0 (0.9) | 30 | 1.8 (0.9) | 106 | 2.1 (0.9) | 0.03† |
| Additional biomarkers |  |  |  |  |  |  |  |  |  |
| Aβ40 | 558 | 158.6 (74.5) | 112 | 166.6 (73.5) | 33 | 168.3 (65.1) | 61 | 146.4 (70.2) | 0.21 |
| Aβ42 | 544 | 7.9 (3.8) | 108 | 8.0 (4.0) | 33 | 8.4 (3.0) | 59 | 7.0 (3.6) | 0.26 |
| t-tau | 449 | 1.7 (0.7) | 75 | 1.7 (0.8) | 20 | 1.8 (0.7) | 68 | 1.7 (0.8) | 0.78 |
| Aβ42/t-tau | 329 | 5.3 (2.7) | 53 | 5.5 (2.7) | 18 | 5.9 (2.3) | 30 | 5.2 (2.6) | 0.52 |
| Aβ42/p-tau181 | 474 | 4.8 (2.9) | 88 | 4.6 (2.7) | 25 | 5.5 (2.9) | 48 | 3.6 (2.6) | 0.004‡ |

\* *p*-values were obtained from the Kruskal-Wallis test.
† After adjusting for multiple comparisons, plasma p-tau181 was significantly higher in the AD group than the CU group.
‡ After adjusting for multiple comparisons, the Aβ42/p-tau181 ratio was significantly lower in the AD group compared to CU or CINAD group.
Abbreviations: CU, cognitively unimpaired; MCI, mild cognitive impairment; CINAD, cognitively impaired but not AD; AD, Alzheimer disease; t-tau, total tau.

**Supplemental Table 2.** Associations between plasma biomarkers and AD-related outcomes.

|  | Biomarker* | AD† |  |  |  | CP with MCI‡ |  |  |  | CP without MCI‡ |  |  |  |
| --- | --- | --- | --- | --- | --- | --- | --- | --- | --- | --- | --- | --- | --- |
|  |  | N | OR | 95% CI | p | N | OR | 95% CI | p | N | OR | 95% CI | p |
| Amyloid biomarkers | Aβ42/Aβ40 | 583 | 0.61 | (0.20 - 2.18) | 0.41 | 721 | 1.81 | (1.01 - 3.23) | 0.05 | 615 | 1.76 | (0.83 - 3.66) | 0.13 |
|  | Aβ40 | 604 | 0.65 | (0.37 - 1.22) | 0.15 | 745 | 0.89 | (0.61 - 1.26) | 0.51 | 636 | 1.17 | (0.75 - 1.77) | 0.48 |
|  | Aβ42 | 589 | 0.69 | (0.34 - 1.46) | 0.31 | 727 | 1.11 | (0.78 - 1.59) | 0.55 | 621 | 1.38 | (0.87 - 2.18) | 0.17 |
| Tau biomarkers | t-tau | 511 | 1.18 | (0.57 - 2.56) | 0.66 | 604 | 1.00 | (0.65 - 1.53) | 0.99 | 531 | 0.82 | (0.46 - 1.41) | 0.47 |
|  | p-tau181 | 751 | 0.77 | (0.39 - 1.51) | 0.44 | 900 | 0.77 | (0.53 - 1.13) | 0.19 | 780 | 0.58 | (0.35 - 0.96) | 0.04 |
| Aβ/Tau | Aβ42/t-tau | 355 | 0.66 | (0.25 - 1.96) | 0.43 | 425 | 1.38 | (0.87 - 2.14) | 0.16 | 373 | 1.57 | (0.83 - 2.90) | 0.16 |
|  | Aβ42/p-tau181 | 513 | 0.86 | (0.39 - 1.96) | 0.72 | 624 | 1.29 | (0.93 - 1.77) | 0.12 | 537 | 1.80 | (1.18 - 2.76) | 0.01 |

\* Biomarkers were log-transformed.
† Models in predicting AD was adjusted for age, age<sup>2</sup>, sex, APOE ε4, time from specimen collection to diagnosis, biomarker batches, research center, and interaction between each biomarker and APOE ε4.
‡ Models in predicting CP was adjusted for age, age<sup>2</sup>, sex, APOE ε2, time from specimen collection to diagnosis, biomarker batches, research center, and interaction between each biomarker and APOE ε2.
Abbreviations: AD, Alzheimer disease; CP, cognitive preservation; MCI, mild cognitive impairment; OR, odds ratio; CI, confidence interval.

**Supplemental Table 3.** Associations between plasma p-tau 181 and AD among APOE ε4 carriers and non-carriers.

|  | Non-Carriers (n = 555) |  |  | Carriers (n = 186) |  |  |
| --- | --- | --- | --- | --- | --- | --- |
|  | OR | 95% CI | *p* | OR | 95% CI | *p* |
| Log-transformed p-tau181 | 0.75 | (0.38 - 1.49) | 0.41 | 6.02 | (2.33 - 17.11) | < 0.001 |
| Age | 1.25 | (0.48 - 4.10) | 0.69 | 1.24 | (0.41 - 4.59) | 0.72 |
| Age<sup>2</sup> | 0.99 | (0.99 - 1.01) | 0.89 | 1.00 | (0.99 - 1.01) | 0.85 |
| Sex | 1.36 | (0.76 - 2.43) | 0.30 | 1.31 | (0.62 - 2.76) | 0.48 |
| Center | 3.24 | (1.75 - 6.19) | < 0.001 | 2.09 | (0.92 - 4.85) | 0.08 |
| Time Diff | 0.50 | (0.20 - 0.97) | 0.08 | 0.77 | (0.35 - 1.18) | 0.40 |

Abbreviations: AD, Alzheimer disease; APOE, apolipoprotein E; OR, odds ratio; CI, confidence interval; Time\_Diff, time from sample collection to diagnosis.

**Supplemental Table 4.** Performance evaluation of plasma biomarkers for AD (distinguishing AD versus CU)

| Model | AUC | Sensitivity | Specificity | PPV | NPV |
| --- | --- | --- | --- | --- | --- |
| Ref* | 0.64 | 0.53 | 0.68 | 0.91 | 0.20 |
| Ref + APOE ε4 | 0.72 | 0.74 | 0.61 | 0.92 | 0.28 |
| Ref + APOE ε4 + individual biomarker | | | | | |
| Aβ42/Aβ40 | 0.81 | 0.63 | 0.88 | 0.98 | 0.21 |
| Aβ40 | 0.80 | 0.68 | 0.82 | 0.97 | 0.22 |
| Aβ42 | 0.81 | 0.68 | 0.88 | 0.98 | 0.23 |
| t-tau | 0.71 | 0.80 | 0.54 | 0.92 | 0.29 |
| p-tau181 | 0.79 | 0.64 | 0.79 | 0.95 | 0.26 |
| Aβ42/tTau | 0.81 | 0.67 | 0.86 | 0.98 | 0.19 |
| Aβ42/pTau | 0.83 | 0.58 | 0.94 | 0.99 | 0.18 |
| Ref + APOE ε4 + multiple biomarkers | | | | | |
| Aβ42/40 + p-tau181 | 0.82 | 0.58 | 0.94 | 0.99 | 0.19 |
| Aβ40 + Aβ42 + Aβ42/40 + t-tau + p-tau181 | 0.83 | 0.72 | 0.83 | 0.98 | 0.20 |

\* The reference model was adjusted for age, age<sup>2</sup>, sex, research center, and time from sample collection to diagnosis.
Abbreviations: AD, Alzheimer disease; CU, cognitively unimpaired; Ref, reference model; AUC, area under the curve;
PPV, positive predictive value; NPV, negative predictive value; APOE, apolipoprotein E.

**Supplemental Table 5.** Performance evaluation of plasma biomarkers for cognitive preservation (distinguishing CU versus CI)

| Model | CP with MCI |  |  |  |  | CP without MCI |  |  |  |  |
| --- | --- | --- | --- | --- | --- | --- | --- | --- | --- | --- |
|  | AUC | Sensitivity | Specificity | PPV | NPV | AUC | Sensitivity | Specificity | PPV | NPV |
| Ref* | 0.63 | 0.49 | 0.70 | 0.41 | 0.77 | 0.64 | 0.48 | 0.74 | 0.29 | 0.87 |
| Ref + APOE ε2 | 0.64 | 0.59 | 0.62 | 0.39 | 0.79 | 0.64 | 0.49 | 0.74 | 0.29 | 0.87 |
| Ref + APOE ε2 + individual biomarker | | | | | | | | | | |
| Aβ42/Aβ40 | 0.69 | 0.62 | 0.67 | 0.41 | 0.83 | 0.74 | 0.83 | 0.55 | 0.24 | 0.95 |
| Aβ40 | 0.68 | 0.66 | 0.63 | 0.40 | 0.83 | 0.73 | 0.72 | 0.63 | 0.25 | 0.93 |
| Aβ42 | 0.69 | 0.65 | 0.62 | 0.39 | 0.83 | 0.74 | 0.77 | 0.60 | 0.24 | 0.94 |
| t-tau | 0.62 | 0.45 | 0.40 | 0.83 | 0.10 | 0.63 | 0.52 | 0.30 | 0.83 | 0.09 |
| p-tau181 | 0.64 | 0.68 | 0.56 | 0.38 | 0.82 | 0.69 | 0.59 | 0.71 | 0.30 | 0.89 |
| Aβ42/t-tau | 0.70 | 0.95 | 0.39 | 0.32 | 0.96 | 0.73 | 0.72 | 0.63 | 0.22 | 0.94 |
| Aβ42/p-tau181 | 0.69 | 0.35 | 0.21 | 0.81 | 0.03 | 0.77 | 0.38 | 0.17 | 0.82 | 0.03 |
| Ref + APOE ε2 + multiple biomarkers | | | | | | | | | | |
| Aβ42/40 + p-tau181 | 0.69 | 0.64 | 0.65 | 0.39 | 0.84 | 0.75 | 0.79 | 0.62 | 0.25 | 0.95 |
| Aβ40 + Aβ42 + Aβ42/40<br>+ t-tau + p-tau181 | 0.71 | 0.84 | 0.49 | 0.33 | 0.91 | 0.77 | 0.88 | 0.60 | 0.23 | 0.97 |

\* The reference model was adjusted for age, age<sup>2</sup>, sex, research center, and time from sample collection to diagnosis.
Abbreviations: CP, cognitive preservation; CI, cognitively impaired; MCI, mild cognitive impairment; Ref, reference model;
AUC, area under the curve; PPV, positive predictive value; NPV, negative predictive value; APOE, apolipoprotein E.

**Supplemental Table 6.**
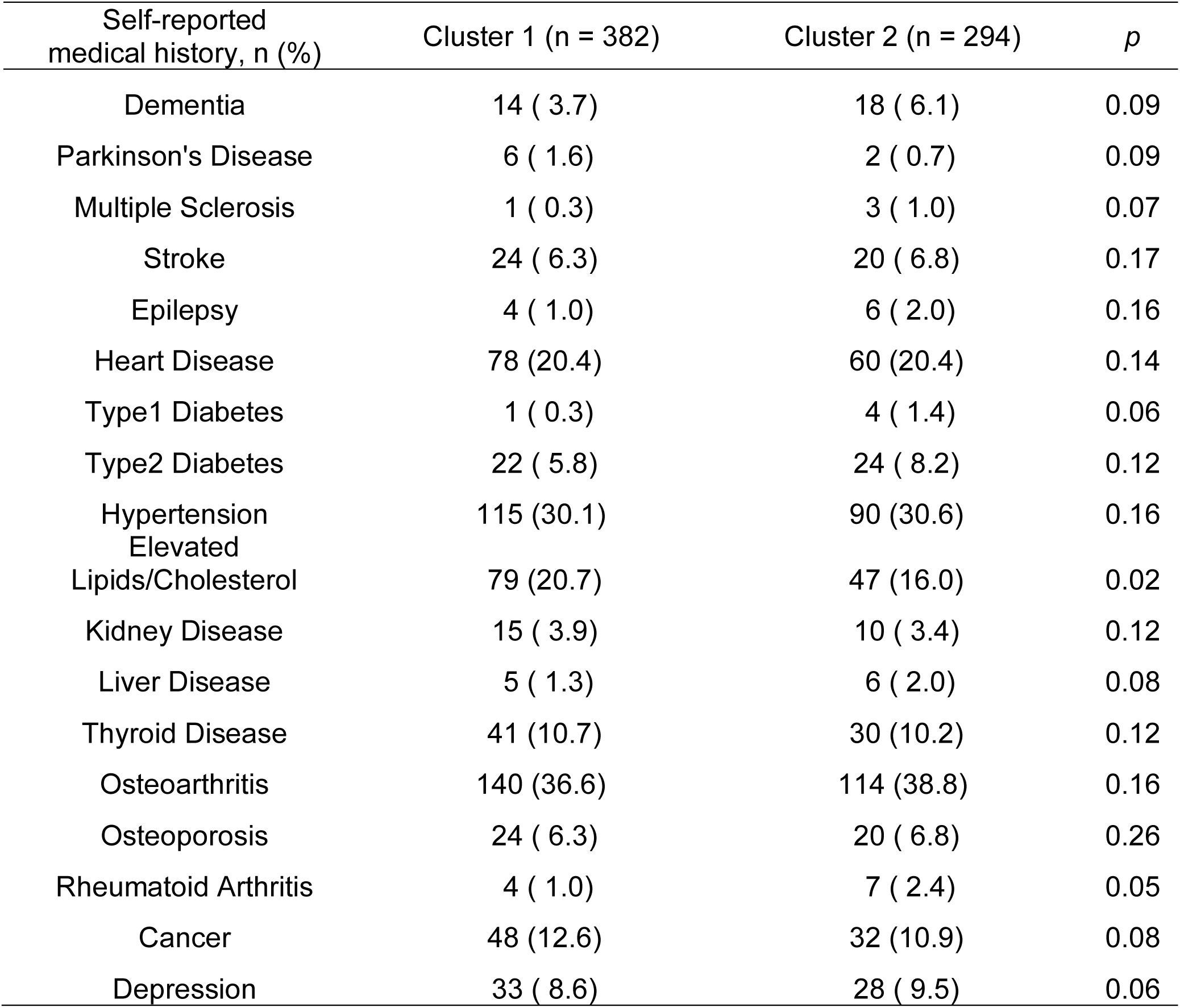
Comparison of medical history between the pre-fixed number of clusters (k = 2)

**Supplemental Table 7.**
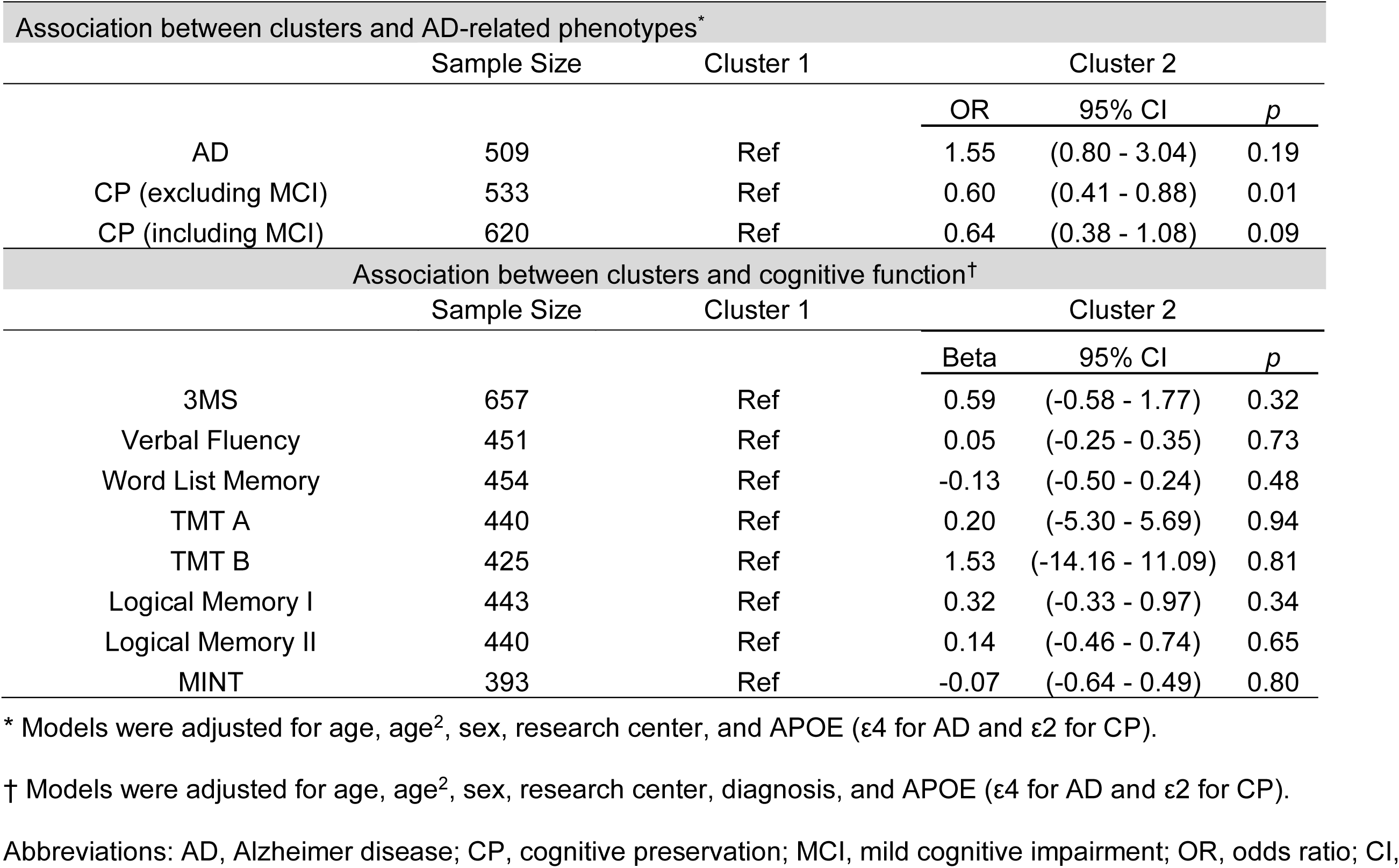
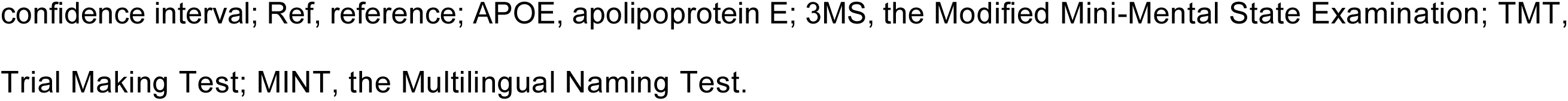
Pre-fixed number of clusters (k = 2) in relationship to AD-related phenotypes and cognitive function.

| Association between clusters and AD-related phenotypes* |  |  |  |  |  |
| --- | --- | --- | --- | --- | --- |
|  | Sample Size | Cluster 1 | Cluster 2 |  |  |
|  |  |  | OR | 95% CI | *p* |
| AD | 509 | Ref | 1.55 | (0.80 - 3.04) | 0.19 |
| CP (excluding MCI) | 533 | Ref | 0.60 | (0.41 - 0.88) | 0.01 |
| CP (including MCI) | 620 | Ref | 0.64 | (0.38 - 1.08) | 0.09 |
| Association between clusters and cognitive function† |  |  |  |  |  |
|  | Sample Size | Cluster 1 | Cluster 2 |  |  |
|  |  |  | Beta | 95% CI | *p* |
| 3MS | 657 | Ref | 0.59 | (-0.58 - 1.77) | 0.32 |
| Verbal Fluency | 451 | Ref | 0.05 | (-0.25 - 0.35) | 0.73 |
| Word List Memory | 454 | Ref | -0.13 | (-0.50 - 0.24) | 0.48 |
| TMT A | 440 | Ref | 0.20 | (-5.30 - 5.69) | 0.94 |
| TMT B | 425 | Ref | 1.53 | (-14.16 - 11.09) | 0.81 |
| Logical Memory I | 443 | Ref | 0.32 | (-0.33 - 0.97) | 0.34 |
| Logical Memory II | 440 | Ref | 0.14 | (-0.46 - 0.74) | 0.65 |
| MINT | 393 | Ref | -0.07 | (-0.64 - 0.49) | 0.80 |

\* Models were adjusted for age, age<sup>2</sup>, sex, research center, and APOE (ε4 for AD and ε2 for CP).
† Models were adjusted for age, age<sup>2</sup>, sex, research center, diagnosis, and APOE (ε4 for AD and ε2 for CP).
Abbreviations: AD, Alzheimer disease; CP, cognitive preservation; MCI, mild cognitive impairment; OR, odds ratio; CI,
confidence interval; Ref, reference; APOE, apolipoprotein E; 3MS, the Modified Mini-Mental State Examination; TMT, Trial Making Test; MINT, the Multilingual Naming Test.

**Supplemental Table 8.**
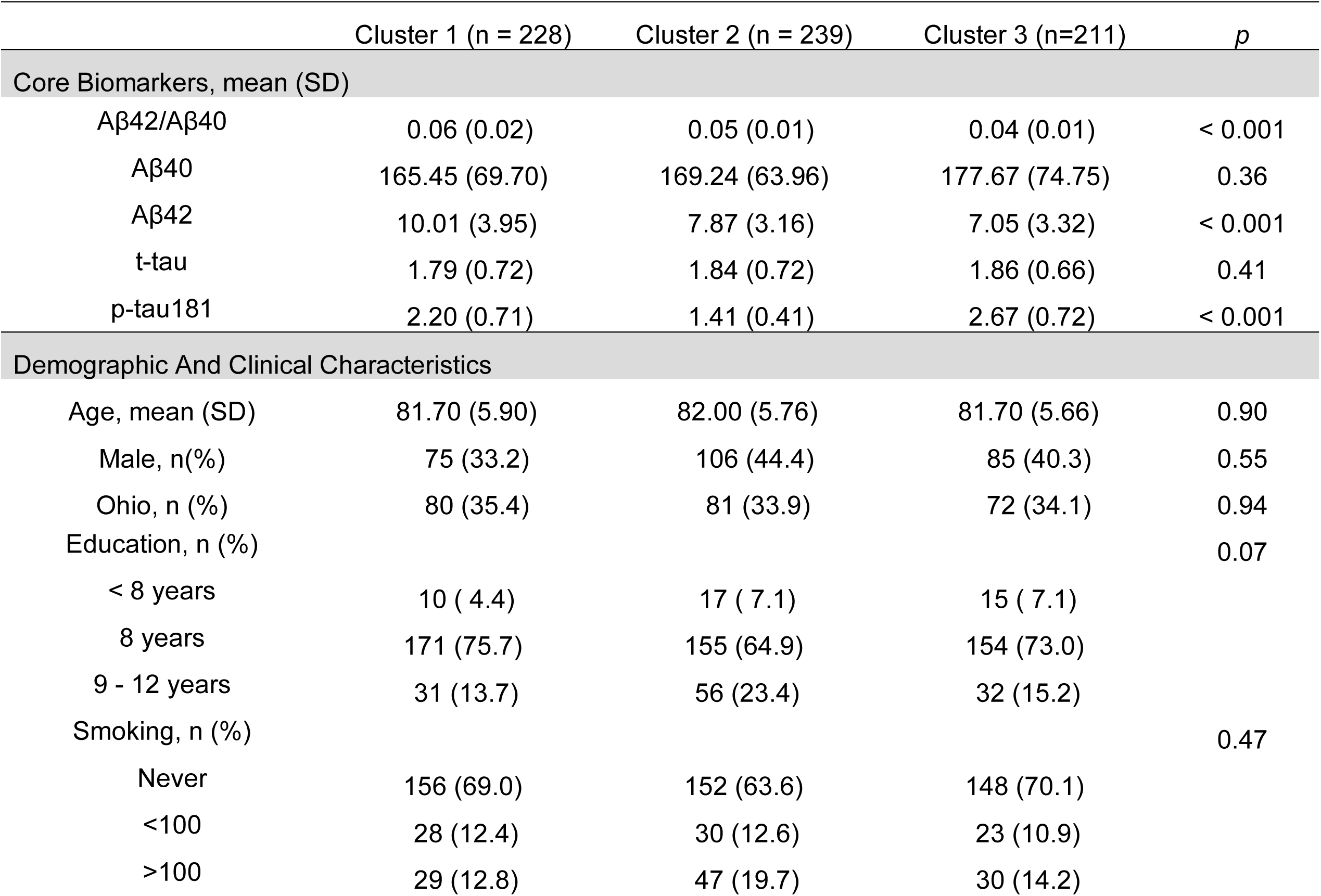
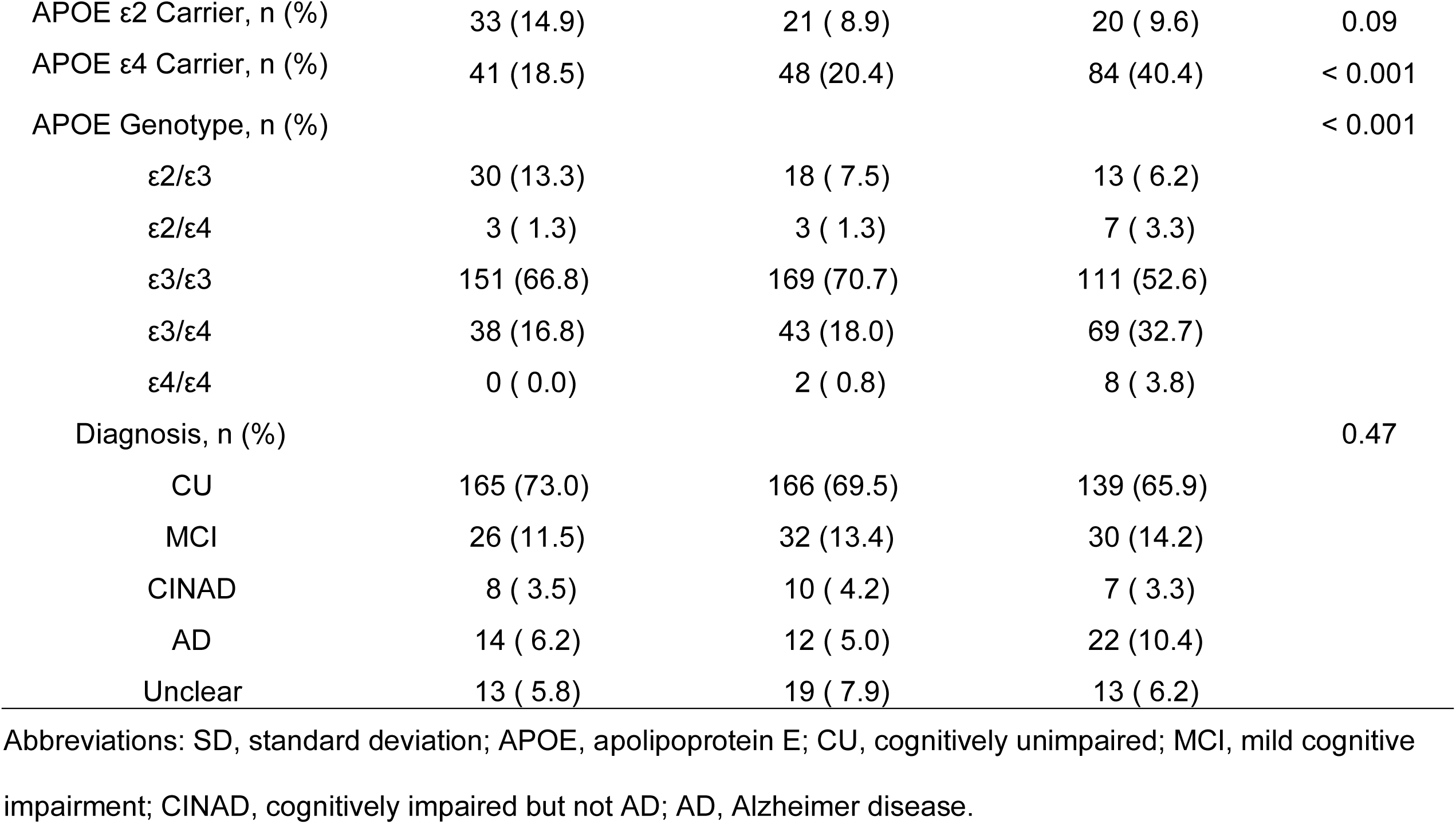
Characteristics of method-derived number of clusters (k = 3) using plasma Aβ42/Aβ40 and p- tau181.

**Supplemental Table 9.** Comparison of medical history between the method-derived number of clusters (k = 3)

| Self-reported medical history, n (%) | Cluster 1 (n = 228) | Cluster 2 (n = 239) | Cluster 3 (n=211) | p |
| --- | --- | --- | --- | --- |
| Dementia | 9 ( 4.0) | 9 ( 3.8) | 14 ( 6.6) | 0.56 |
| Parkinson's Disease | 3 ( 1.3) | 4 ( 1.7) | 1 ( 0.5) | 0.35 |
| Multiple Sclerosis | 0 ( 0.0) | 2 ( 0.8) | 2 ( 0.9) | 0.41 |
| Stroke | 13 ( 5.8) | 14 ( 5.9) | 17 ( 8.1) | 0.69 |
| Epilepsy | 1 ( 0.4) | 4 ( 1.7) | 5 ( 2.4) | 0.28 |
| Heart Disease | 45 (19.9) | 50 (20.9) | 43 (20.4) | 0.24 |
| Type1 Diabetes | 1 ( 0.4) | 1 ( 0.4) | 3 ( 1.4) | 0.45 |
| Type2 Diabetes | 10 ( 4.4) | 17 ( 7.1) | 19 ( 9.0) | 0.19 |
| Hypertension | 68 (30.1) | 70 (29.3) | 67 (31.8) | 0.69 |
| Elevated Lipids/Cholesterol | 50 (22.1) | 45 (18.8) | 31 (14.7) | 0.12 |
| Kidney Disease | 11 ( 4.9) | 7 ( 2.9) | 7 ( 3.3) | 0.51 |
| Liver Disease | 6 ( 2.7) | 0 ( 0.0) | 5 ( 2.4) | 0.10 |
| Thyroid Disease | 29 (12.8) | 23 ( 9.6) | 19 ( 9.0) | 0.40 |
| Osteoarthritis | 84 (37.2) | 87 (36.4) | 83 (39.3) | 0.70 |
| Osteoporosis | 22 ( 9.7) | 11 ( 4.6) | 11 ( 5.2) | 0.07 |
| Rheumatoid Arthritis | 3 ( 1.3) | 4 ( 1.7) | 4 ( 1.9) | 0.53 |
| Cancer | 28 (12.4) | 29 (12.1) | 23 (10.9) | 0.79 |
| Depression | 10 ( 4.4) | 29 (12.1) | 22 (10.4) | 0.01 |

**Supplemental Table 10.**
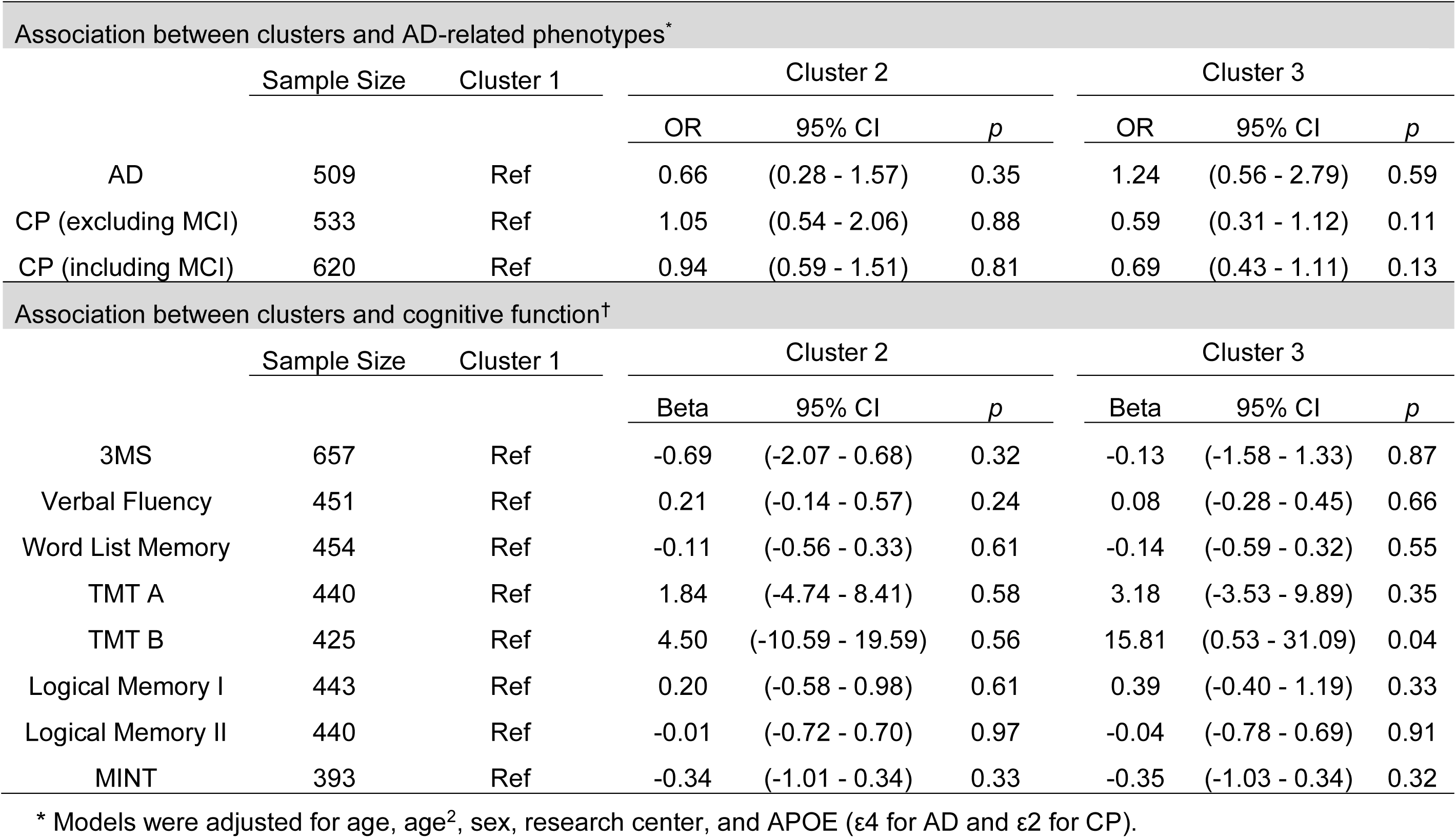
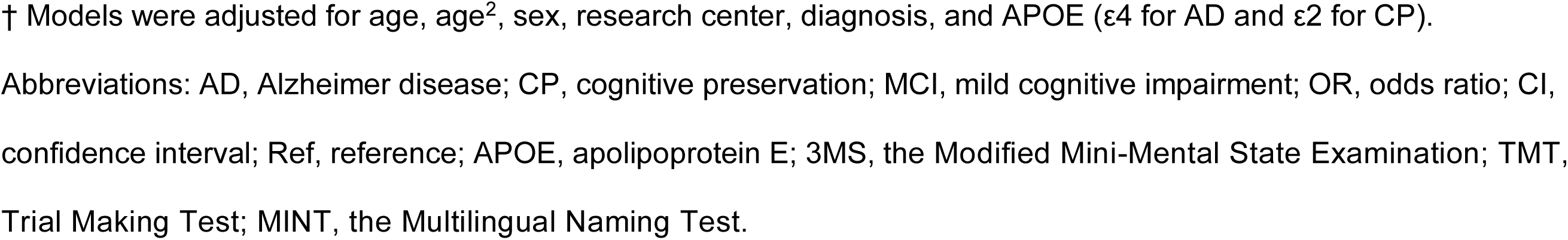
Method-derived number of clusters (k = 3) in relationship to AD-related phenotypes and cognitive function.

| Association between clusters and AD-related phenotypes* |  |  |  |  |  |  |  |  |
| --- | --- | --- | --- | --- | --- | --- | --- | --- |
|  | Sample Size | Cluster 1 | Cluster 2 |  |  | Cluster 3 |  |  |
|  |  |  | OR | 95% CI | *p* | OR | 95% CI | *p* |
| AD | 509 | Ref | 0.66 | (0.28 - 1.57) | 0.35 | 1.24 | (0.56 - 2.79) | 0.59 |
| CP (excluding MCI) | 533 | Ref | 1.05 | (0.54 - 2.06) | 0.88 | 0.59 | (0.31 - 1.12) | 0.11 |
| CP (including MCI) | 620 | Ref | 0.94 | (0.59 - 1.51) | 0.81 | 0.69 | (0.43 - 1.11) | 0.13 |
| Association between clusters and cognitive function† |  |  |  |  |  |  |  |  |
|  | Sample Size | Cluster 1 | Cluster 2 |  |  | Cluster 3 |  |  |
|  |  |  | Beta | 95% CI | *p* | Beta | 95% CI | *p* |
| 3MS | 657 | Ref | -0.69 | (-2.07 - 0.68) | 0.32 | -0.13 | (-1.58 - 1.33) | 0.87 |
| Verbal Fluency | 451 | Ref | 0.21 | (-0.14 - 0.57) | 0.24 | 0.08 | (-0.28 - 0.45) | 0.66 |
| Word List Memory | 454 | Ref | -0.11 | (-0.56 - 0.33) | 0.61 | -0.14 | (-0.59 - 0.32) | 0.55 |
| TMT A | 440 | Ref | 1.84 | (-4.74 - 8.41) | 0.58 | 3.18 | (-3.53 - 9.89) | 0.35 |
| TMT B | 425 | Ref | 4.50 | (-10.59 - 19.59) | 0.56 | 15.81 | (0.53 - 31.09) | 0.04 |
| Logical Memory I | 443 | Ref | 0.20 | (-0.58 - 0.98) | 0.61 | 0.39 | (-0.40 - 1.19) | 0.33 |
| Logical Memory II | 440 | Ref | -0.01 | (-0.72 - 0.70) | 0.97 | -0.04 | (-0.78 - 0.69) | 0.91 |
| MINT | 393 | Ref | -0.34 | (-1.01 - 0.34) | 0.33 | -0.35 | (-1.03 - 0.34) | 0.32 |

\* Models were adjusted for age, age<sup>2</sup>, sex, research center, and APOE (ε4 for AD and ε2 for CP).
† Models were adjusted for age, age<sup>2</sup>, sex, research center, diagnosis, and APOE (ε4 for AD and ε2 for CP).
Abbreviations: AD, Alzheimer disease; CP, cognitive preservation; MCI, mild cognitive impairment; OR, odds ratio; CI, confidence interval; Ref, reference; APOE, apolipoprotein E; 3MS, the Modified Mini-Mental State Examination; TMT, Trial Making Test; MINT, the Multilingual Naming Test.

